# COVID-19 and Health System Response for Management of Diabetes in Bangladesh: A national qualitative study of patients with diabetes and key healthcare professionals/policy makers

**DOI:** 10.1101/2022.12.18.22283405

**Authors:** Shamim Hayder Talukder, Tasneem Islam, Kazi Fayzus Salahin, Shahin Akter, Dina Farhana, Ummay Farihin Sultana, Sheikh Mohammed Shariful Islam, Zumin Shi, Akhtar Hussain, Brian Godman, Amanj Kurdi

## Abstract

**Background:** Uncontrolled diabetes can lead to exacerbation of symptoms and life-threatening complications. Consequently, there is a need to explore patient experience regarding the prevention and treatment of diabetic patients amidst the restrictions and lockdown measures in response to COVID-19. The objective of this study was to assess the response of the healthcare system for preventive care and treatment of people with diabetes in Bangladesh during COVID-19, and to analyze the health-seeking behavior of diabetes patients amidst social distancing and lockdown measures

**Method:** A descriptive qualitative design was used to collect data regarding the ability of people living with diabetes to access medication, laboratory services, and preventative care during the pandemic. The data collection process involved 12 focus group discussions (FGDs) with people living with diabetes, and 30 key informants’ interviews (KIIs) with senior diabetologist, health service managers, leaders of different diabetes-related associations, and policymakers from the local to the national level. The discussion issues were structured around the WHO framework that describes health systems in terms of six “building blocks”. In addition, different treatment guidelines, scientific articles, relevant reports, and 20 well-circulated newspapers were analyzed concerning the treatment of diabetic patients.

**Results:** 44% of the respondents were aged 55-60 years, with an informed noticeable disruption of essential diabetes care services, intensified by high COVID-19 infection rates. Besides, 78% of the service receiver participants reported not seeing any government-issued public announcements regarding diabetes management on television or newspapers. There are also concerns with the current heath sector.

**Conclusions:** The study findings highlighted major concerns surrounding the healthcare response to deliver care for patients with diabetes during the pandemic, driven mainly by restricted access to treatment under lockdown measures coupled with a reluctance from health care providers to see patients due to high COVID-19 infection rates arising from concerns with a lack of personal protective equipment. Necessary measures can gradually bring some change in the healthcare system promote healthy lifestyles and adherence to prescribed medicines together with raising awareness about the potential risk factors of diabetes.

## Introductions

Bangladesh is a developing country with considerable economic growth, with currently a population of 165 million [1] . Alongside this, there has been rapid urbanization for the past several decades [2]. Such process of development and urbanization contributes to the rise in the burden of type 2 diabetes, exacerbated by the popularity of processed food, irregular mealtimes, and reduced physical activity [3]. As a result, the prevalence of diabetes is comparatively higher in urban areas versus rural areas in Bangladesh, with one in every 10 adults now having diabetes in Bangladesh [4]. The speared of diabetes is mostly seen in Dhaka and Chittagong, the two extensive cities of Bangladesh [5]. Diabetes increases steeply with increasing BMI for both genders, disproportionately affecting patients in these two densely populated cities [6]. They are also the locations with most positive COVID-19 cases in Bangladesh [7]. Overall, the prevalence of diabetes among both women and men above 35 years in Bangladesh increased similarly between 2011 and 2017 from 11% to 14%, indicating the need for expanded diabetes awareness programs as well as programmes to improve the care of patients with diabetes to reduce complication rates with their considerable impact on morbidity, mortality and costs [8]. This is because there are concerns with the identification of patients with Type 2 diabetes in the first place as well as concerns whether they are optimally managed thereafter to prevent subsequent complications including cardiovascular disease [9].

COVID-19 emerged as a rapidly evolving pandemic, impacting nearly 220 countries and more than 650,329,275patients worldwide, with more than 6,647,714 deaths as of December 06, 2022 [10]. Till late November, 2022 Bangladesh has identified 2,036,622 COVID-19 cases among them death cases are 29,434 and about 80% of the deaths occurred above 50 years of age [7]. The rapid implementation of preventative measures helped to slow the spread of the virus, and its impact, similar to a number of other Asian countries [11,12]. Known risk factors for severe cases of COVID-19 including DM, COPD, CVD and HTN (Hypertension) have a higher prevalence in low- and middle-income countries (LMICs) including Bangladesh, and these conditions are generally more poorly controlled in LMICs. Consequently, patients with COVID-19 in these countries at greater risk of complications and their associated impact on morbidity, mortality and costs [8,13,14]. However, a rapid assessment by the WHO found that 75% of countries reported interruptions to NCD services during the recent pandemic [15,16]. A recent study conducted in LMICs among 734 hospitalized patients diagnosed with COVID-19 found that 19.8% had diabetes and the frequency of patients requiring insulin increased three-fold [17]. This has important implications regarding the management of patients with diabetes during pandemics especially where there are concerns with access to insulins and their affordability, which are key issues among LMICs [18,19]. In addition, concerns with the identification and adequate management of patients with Type 2 diabetes during the current pandemic with lockdown and other measures [20].

Consequently, with the growing burden of diabetes in Bangladesh [9], there is a need to explore the health system management components’ response to the treatment of patients with diabetes during the current COVID-19 outbreak in Bangladesh. Across Bangladesh, approximately eight million diabetic patients visited diabetic centers to receive routine care with regular follow-up before the first confirmed case of COVID-19 [21]. After the government implemented initial lockdown measures in March 2020 until May 2020, all health service centers became stagnant with many health service providers becoming infected with COVID-19 [22,23]. Lockdown measures also reduced the means of physical exercise as people were limited to the confines of their homes in addition to reduced access to healthcare services [9]. There was also a scarcity of medicines at that time and their inaccessibility [24]. Many patients also began utilizing mobile health services for their diabetic management [25]. However, we are unaware of a comprehensive robust study to fully explore and understand the health system response to the management of patients with diabetes during COVID-19 in Bangladesh. This in important as it will provide key learning lessons necessary to improve future care in a high diabetic prevalence country building on the findings of pilot and other studies undertaken in Bangladesh. Consequently, the objective of this study was to assess the response of the healthcare system for preventive care and treatment of people with patients with diabetes in Bangladesh during COVID-19, and to analyze the health-seeking behavior of these patients amidst social distancing and lockdown measures to provide future direction.

## Methodology

### Study Design and Participant

A descriptive design was undertaken that was multifaceted. This included, firstly, a scoping literature review, which was followed by qualitative research involving focus group discussions (FGD) with patients with diabetes and key informant interviews (KII) with health service providers from different hospitals and diabetes care centers at national and local level in Bangladesh.

### Recruitment Strategy

The sample size for the qualitative research was determined purposively to provide the maximum variations in terms of patients’ demographics. The data collection process involved 30 KIIs with senior diabetologist, health service managers, leaders of different diabetes-related associations, and policymakers. They were communicated though email and scheduled for the KII. The policymaker refers to a person who is responsible for, or involved in, formulating policies and have a voice in changing policy. Besides, 12 FGDs were conducted with both the male and female patients living with diabetes and healthcare service providers. A, snowball sampling technique was used to identify people who have diabetes for at least for five years with a minimum age of 18 for the FGDs.The healthcare service providers were scheduled and recruited through email.

### Conduction of KII and FGD

Interviews with KIIs were performed through zoom and the participants were selected purposively among those who were available during the study period. The timeframe for each FGD was approximately one and a half hours. All KIIs and FGDs were conducted by qualitative data collectors through telephone and Zoom calls due to countywide lockdown in 2021. All KIIs and FGDs were facilitated by the researchers themselves.

### Consent of Participant

The study participation was completely voluntary. The KIIs and FGDs were conducted by using online platforms including zoom, google meet as per respondents’ user convenience and verbal consent was obtained before moving forward with the questionnaire. Individual information was preserved to maintain the confidentiality as per IRB.

### Discussion points and Data Management

A separate KII checklist, and FGD guideline was used for the study with the checklist and guideline designed considering specific probes on key discussion issues (**Error! Reference source n ot found**.). The specific probes were identified by inter-group discussion among the co-authors and rapid brainstorming, centering on the discussion issues following a thorough literature review.

The literature review included diabetes management guidelines, developed by Director General of Health Service (DGHS) under the Ministry of Health and Family Welfare of Bangladesh, Diabetes Association of Bangladesh (DAB) and World Health Organization (WHO); COVID-19 treatment guidelines, scientific articles, different reports, and newspaper entries. For the scientific articles, different search engines were used including Google.com, Google Scholar, EndNote, and searched using keywords including Diabetes, COVID-19, Healthcare Management, Diabetes Treatment Guidelines, COVID-19 Treatment Guidelines, Health System Response. In addition, twenty (20) high circulated online newspapers available in Bangladesh (10 in English and 10 in Bengali) were reviewed with the same key words from July to August 2020.

The findings were developed into specific probes which were structured around the WHO framework that describes health systems in terms of six “building blocks”, i.e.: (1) service delivery, (2) health workforce, (3) health information systems, (4) access to essential medicines, (5) financing, and (6) leadership/governance [95] (Table 1). However, not all the questions were relevant for both people living with diabetes and physicians.

**Table 1.**
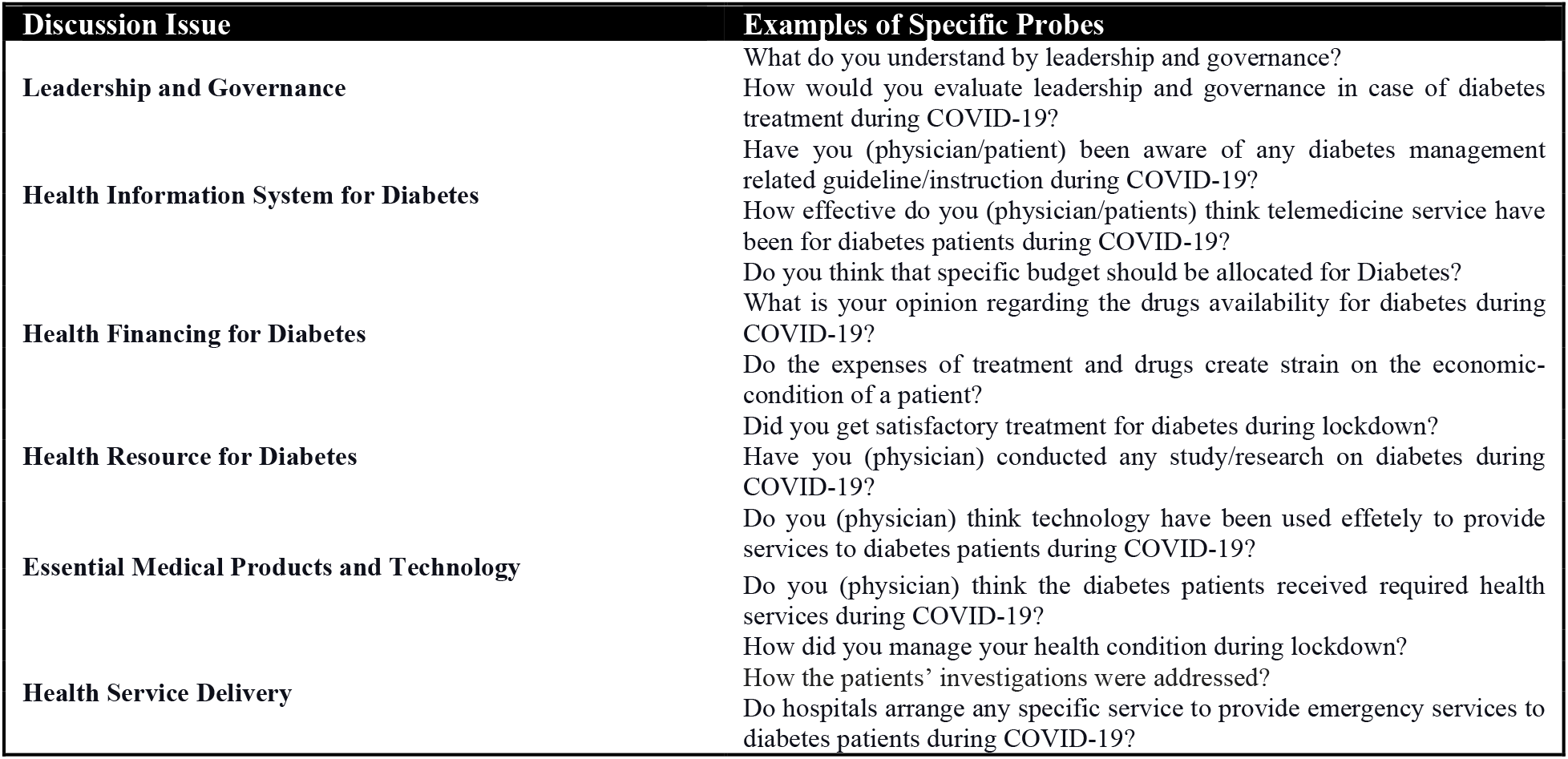
Study Tools Issues

The checklists and guideline were subsequently field tested and the feedback received were incorporated before finalization and utilization during data collection from the target respondents.

After conducting the KIIs and FGDs, the data were transcribed for analysis, and then the transcribed reports were translated back and forth from Bengali to English until each translation became sufficiently similar to the previous language to ensure an accurate translation.

### Data Analysis

The transcripts were analyzed using qualitative content analysis technique [26,27]. Transcripts of both KIIs and FGDs were divided into the themes. These were Healthcare Service and Access to Drugs, including two respondents’ categories that are people living with diabetes and health service provider. The analysis was performed by coding the emerging sub-themes and presenting them under wider themes based on the six components of health systems including leadership and governance, health information systems, health financing, health resources, essential medical products and technology, and health service delivery. Researchers conducted the analysis manually, following the logical framework of the study to achieve the objectives.

Further analysis was undertaken under the six pre-determined components of health system. With respect to the analysis, each transcript was read several times, and irrelevant parts removed. The study employed each primary question as an overarching category (deductive coding). These broad categories were further broken down into subcategories based on the findings that emerged from analyses of the transcripts (inductive coding). Answers to a question bearing the same meaning and content were assembled and ascribed a common code. Initially, the researchers analyzed all the transcripts to generate an initial series of codes for each of the six primary questions. Subsequently, the researchers independently coded all the data which came from literature review, KIIs, and FGDs. Afterwards, all these data have been transcribed. The results were presented as per the study-specified category, grounding on the treatment mode of PLWD and socio-demography of respondents. The result of analysis was discussed during consensus meetings, and any differences in interpretation were resolved at this time. After that, the second round of analysis was independently undertaken by the team leader and other researchers (KFS and UFS); wherein all the ascribed codes were again reviewed, followed by a final discussion among the researchers of the study.

## Results of the Study

### Socio-demography of the Respondents

A total of 126 urban participants participated in the study, and among them poor slum people living with diabetes were involved. Twelve diabetes patients’ groups (96 in total) were select for FGDs, eight participants in each. Among them more than 50% were male. Among all the study respondents, the average ages of the were 52.08 years (SD±13.02) (Table 2). 44% of the respondents were in the age group 41-50 years. However, 48% of the respondent were highly educated (post-Graduate). Among them, 52% were working in different private institutions. Furthermore, 47% of the respondents were in “Middle Income” group in the socio-economies context. With respect to religious beliefs, 80% were Muslim and 20% were of other religion, with the majority of patients married (83%). (Table 2). Most patients were taking oral medicines with only a minority on insulin (20%).

**Table 2.**
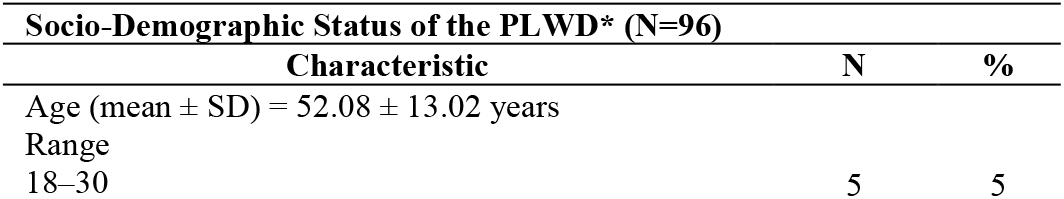
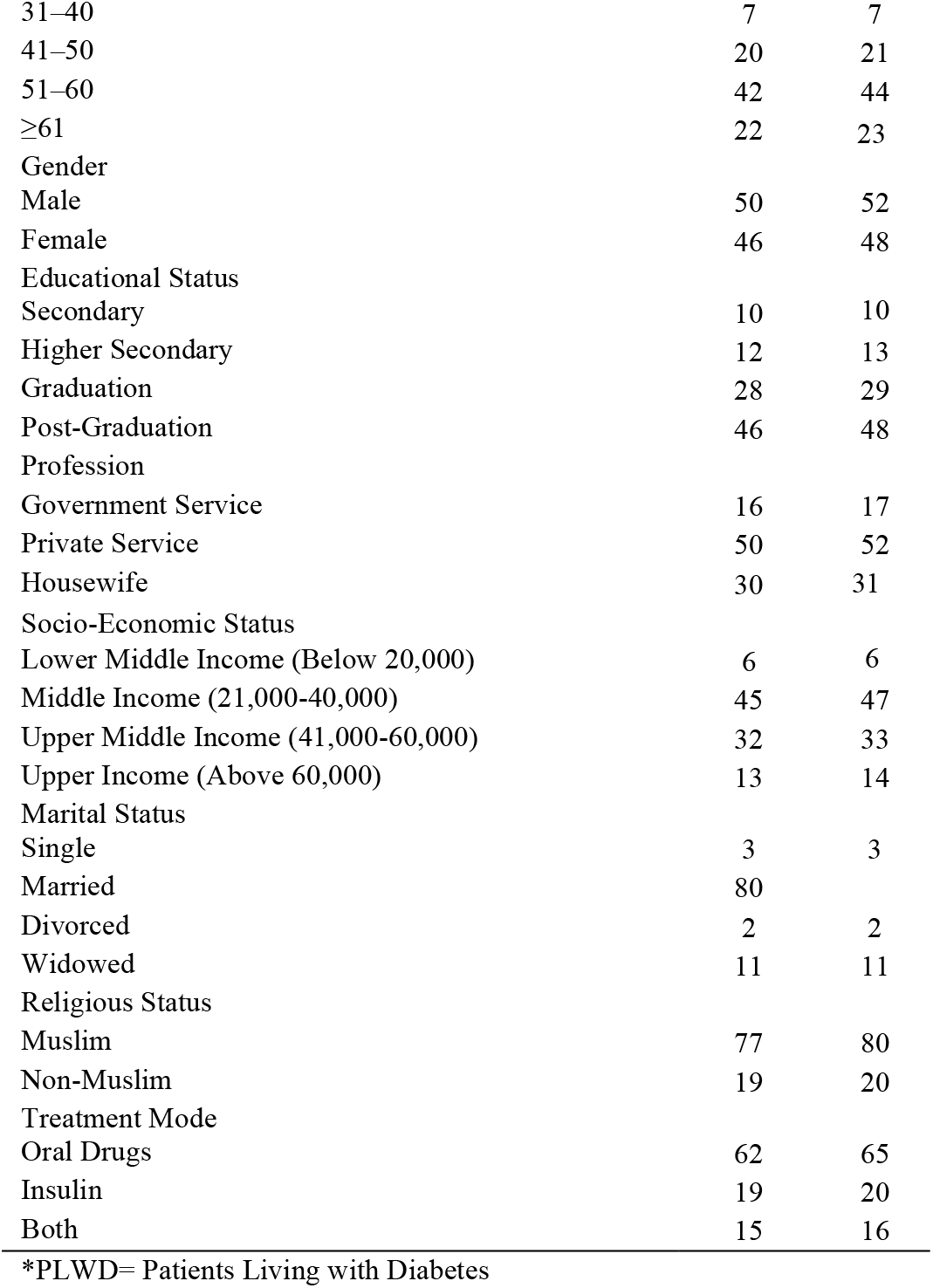
Socio-Demography of the PLWD

From the service providers and other stakeholders (N=30), the mean age was 55.33 years (SD±9.31), with 62% male and the majority physicians (40%) (Table 3).

**Table 3.**
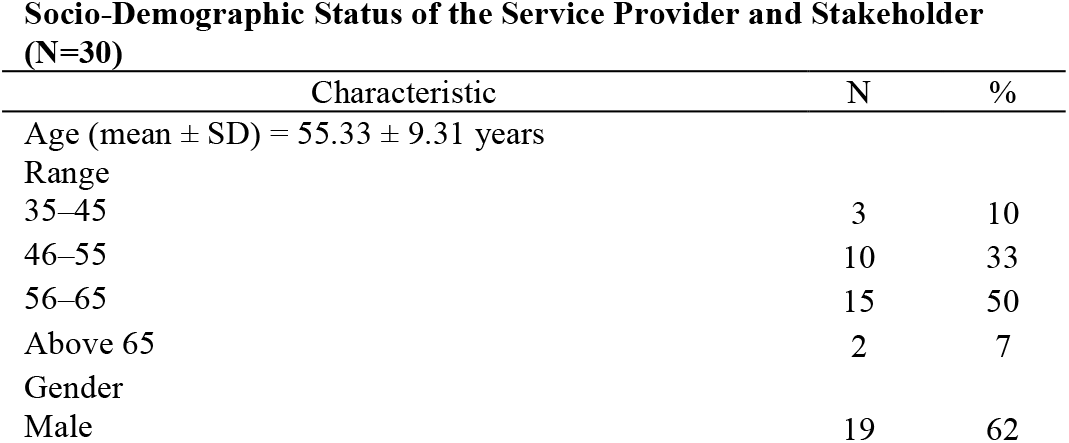
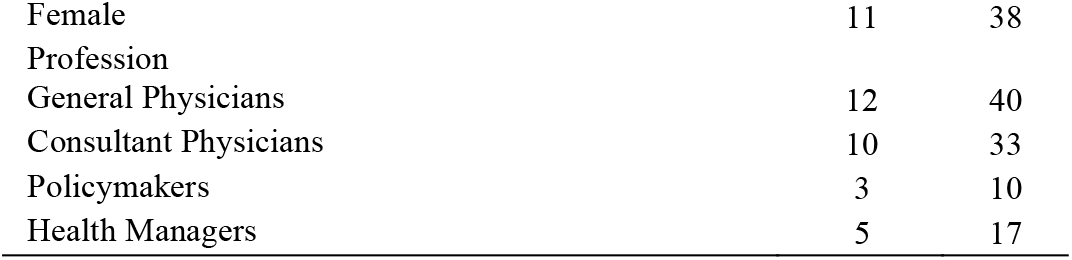
Socio-Demography of the Health Service Provider and Key Stakeholder

The response of the participants was mostly grounded on the theme named health service delivery and access to drug [Table 4], considering the significant challenges in providing essential diabetes care services in

**Table 4.**
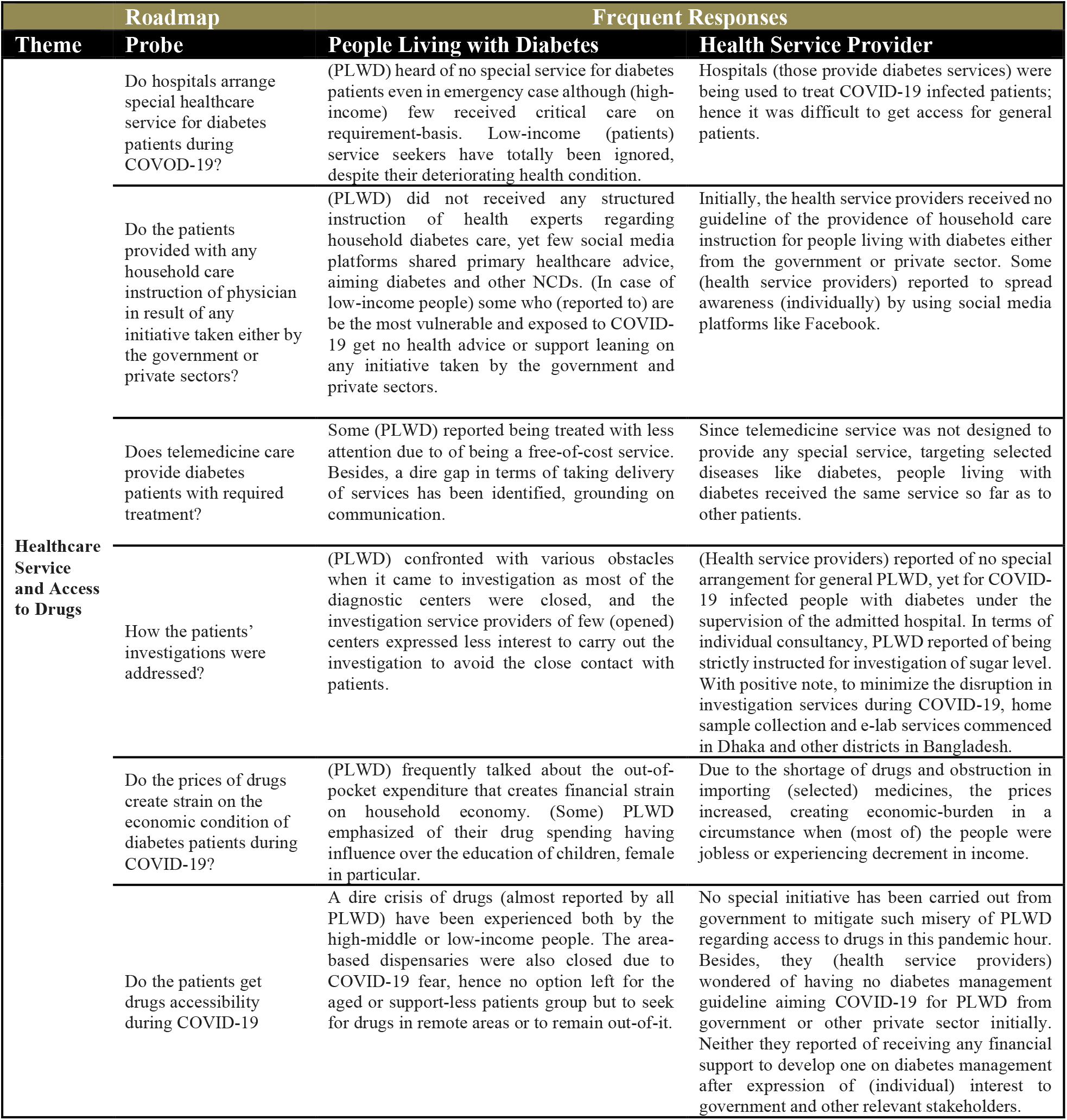
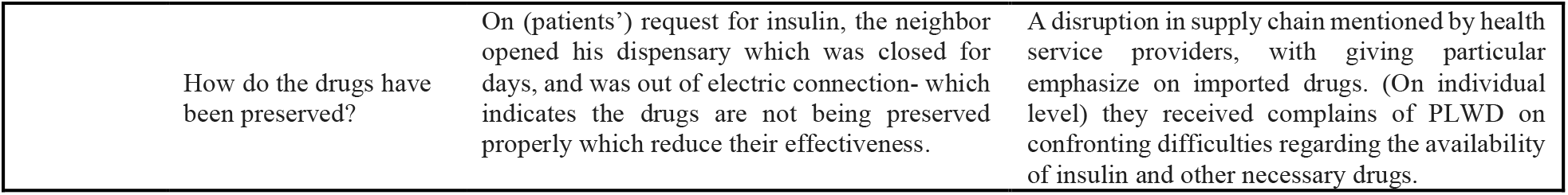
Themes and Response of the Participants

## Health Service Response for Diabetes

### Leadership and Governance for Diabetes

#### Findings from Interviews and FGDs

Most of the respondents (78%) from FGD and KII show minimum satisfaction towards the leadership and governance for diabetes care in Bangladesh during COVID-19 [Table 5; Table 6]. Whilst a guideline for diabetic patients was published in April 2020, a large number of health service providers and receivers reported they were unaware of its existence. As a result, they experienced a lack of guidance from the responsible authorities during the initial lockdown period in April and May 2022 [6,7,10,15,28,28–94]. During lockdown, most of the healthcare professionals (HCPs) were engaged and there was an acute shortage of personal protective equipment (PPE), resulting in little or no opportunity for specialists to treat or attend patients in the various diabetic healthcare centers or clinics [Table 4]. A number of patients also reported during the FDG of having incomplete instructions when consulting with family doctors. One thing that nearly all of the respondents reasoned was that the actual rate of COVID-19 infection among diabetic patients might be higher than the reported data in various news or communication channels [6,15,34,36,43,47–50,52–54,56–58,58–63,65–68,70,71,74–78]

**Table 5.** Interview Key Finding on the Opinion on Diabetes Management during COVID-19 Frequent Responses of PLWD

| Theme | Sub-Thematic Area | Probe | People Living with Diabetes |
| --- | --- | --- | --- |
| Healthcare Service and Access to Drugs | Understanding of leadership and governance | What do you understand by leadership and governance? | A process maintained by government to lead health related issues within the country (as per the utterance of PLWD). |
|  |  | How would you evaluate leadership and governance during COVID-19? | Initially confrontation of confusion regarding the management of diabetes reported due to having no instruction or guideline for individual-level measure. |
|  | Health Information System for Diabetes | Have you been aware of diabetes management related guideline during COVID-19? | On my request my daughter searched for if any instruction have been shared in TV or social media for diabetes patients, found nothing- it was difficult to decide the dos and don'ts. |
|  |  | How effective do you (physician) think telemedicine service have been during COVID-19? | PLWD reported of dire communication gap regarding accessing /receiving services, with particular emphasize on insufficient and less-skilled human resource. |
|  | Health Financing for Diabetes | Do you think that specific budget should be allocated for Diabetes? | PLWD from low economic setting strongly think of specific allocation of budget will benefit them. |
|  |  | What is your opinion regarding the drugs availability for diabetes during COVID-19? | Countrywide lockdown highly impacted the availability of drugs: inconvenience of the closure of dispensaries nearly mentioned by every PLWD. In terms of imported drugs, a sudden raise of price made insulin unaffordable for PLWD in low-economic setting. |
|  | Health Resource for Diabetes | Did you get satisfactory treatment for diabetes during lockdown? | PLWD expressed discontentment regarding physically availing diabetes care services during lockdown. They barely mentioned of arrangement for diabetes care from government. (PLWD reported) to receive less attention from physician in telemedicine service due to its being free of cost. |
|  | Essential Medical Products and Technology | Do you (physician) think technology have been used effectively to provide services to diabetes patients during COVID-19? | PLWD were concerned of the telemedicine service, emphasizing on the failure regarding the documentation and analyze of historical data of them. Besides, most of them claimed not to understand the prescription given over call; which indicates to the gap created due to not producing electronic prescription. |
|  | Health Service Delivery | How did you manage your health condition during lockdown? | For lockdown the daily-walk of PLWD have been hampered, hence the front premises or roof top have been used (by mostly), the traditional food seeking behavior (herbal ingredients and daily use of plants like Orjun, Nim etc) raised in response to getting less access to marketized food. Yet, there was no substitute for medicines PLWD takes on a regular basis, even (some reported) purchased insulin from dispensaries that were shut-down for days, which indicates that the insulin was not kept in required temperature. |

**Table 6.**
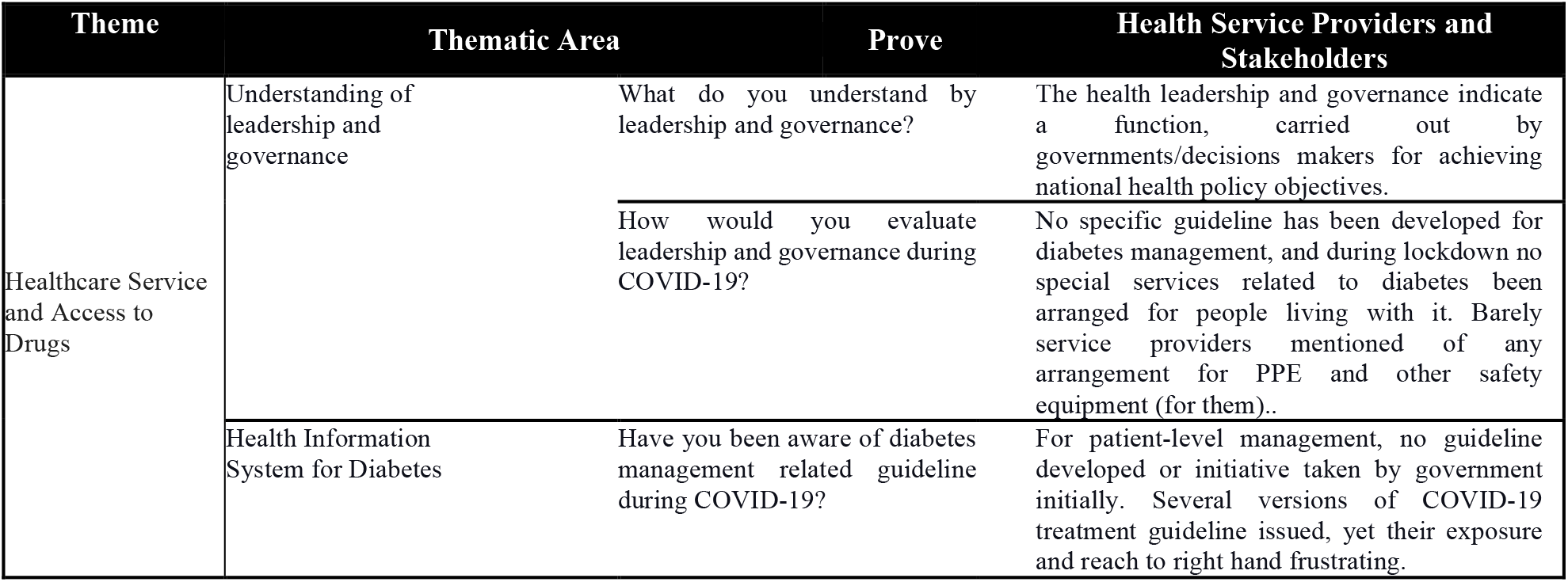
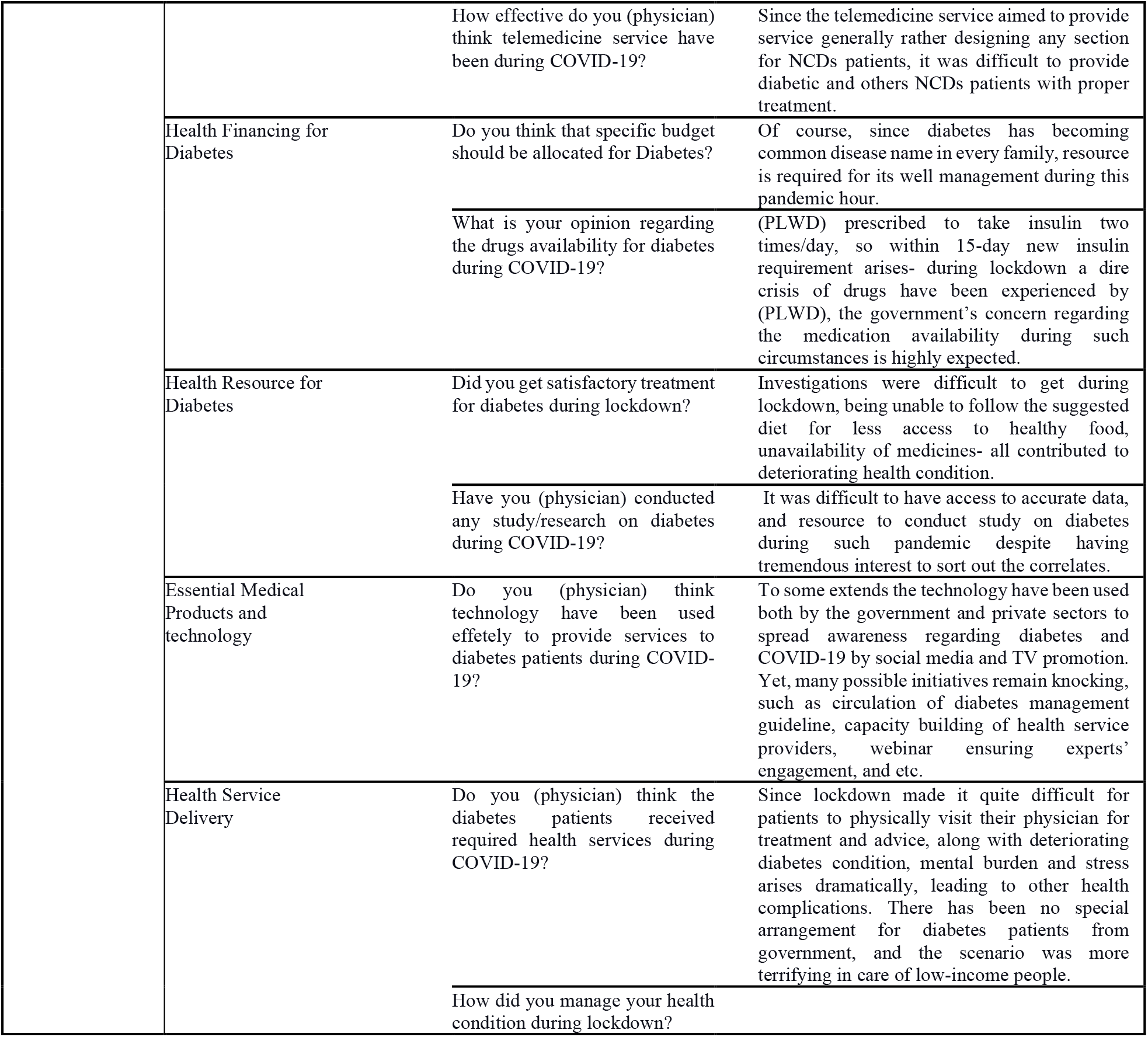
Interview Key Finding on the Opinion on Diabetes Management during COVID-19 Frequent Responses of Service Providers and Stakeholders

When the service providers were asked whether they received or were aware of any diabetes treatment or service guidelines regarding COVID-19, most of the general and consultant physicians were unaware.

> “*A fright was developed inside as COVID-19 was all new then, I seek for guidance first but the sacrifice of patients’ lives made me go without it*” …(p6)

Experts determined that the data needed for proper epidemiological analysis was severely lacking due to the lack of comprehensive testing [95]. This follows similar complaints of many patients dying with COVID-like symptoms, but no way of confirmation [95].

The government established a national technical committee for COVID-19 to provide technical support for planning and managing health services throughout the country. This committee consisted of 21 members among whom only one member’s expertise included NCDs. Most of the respondents of KII shed light on the government’s development of a clinical management guideline for COVID-19, which was updated eight times, but diabetes management was not addressed within the first few versions.

> *“As a doctor, it shocked me that most of the COVID-19 aimed guidelines allocated a very little ground for NCDs while people living with it are the most vulnerable one”* …*(p12)*

Moreover, many service providers thought the public healthcare system lacked an adequate emphasis on physician and frontline healthcare workers’ safety regarding diabetes management [Table 6]. The Diabetic Association Network (DAN) centers were unable to act upon proper guidance to manage their patients. On an individual level, many groups attempted to share guidelines in social media or television announcements, which seem to have a positive impact on the public as they often mentioned these as helpful during the FGDs. Patients highly appreciated some of the individual physician’s leadership, informing the public regarding COVID-19 and key safety issues.

#### Findings from Literature Search

During study period, 11 newspaper reports indicated no new deaths caused by COVID-19 until the end of March. However, there was very low (<1%) COVID-19 diagnosis testing and patients monitoring for COVID-19. These newspaper reports also revealed that many physicians refused to see patients without having a COVID-19 test report . However, the study finds that the shortage of testing kits and restrictive COVID-19 diagnosis policies made it difficult for those diabetes patients who were given investigations that have been needed to be seen by their healthcare providers. Majority news reports reflect a lack in the regulation from the higher authority on how and who should be treated amongst the pandemic, which not only alarmed healthcare providers, rather hindered the possibilities of providing health care services as well.

#### Health Information System for Diabetes

##### Findings from Interviews and FGDs

The FGDs showed that, public awareness towards diabetes was limited in the past six months. 78% of the service receiver participants reported not seeing any government-issued public announcements regarding diabetes management on television and newspapers [Table 4]. Whilst the government created an online telemedicine service platform to reach patients from remote locations, 82% of the service provider participants on this telemedicine line responded not to be explicitly prepared regarding the management of diabetic patients, which led to further inadequate services and decreased the trust between diabetic patients and telemedicine healthcare services [Table 4; Table 5].

However, almost all the participants, both patients and service providers, mentioned an appreciable communication gap in terms of providing and receiving telemedicine service.

> *“It was my first-time telemedicine service experience, I found it difficult to make understand my condition and complications neither understood the physicians’ advice properly” … (FDG-ph-p4)*
>
> *“I prescribed the patient for high blood pressure, cough and fever after a complex explanation of condition, but immediately before cutting the call, she added of having diabetes which turned the table” …(FGD-HCP-p21)*

Participants who treated COVID-19 patients stated that the most complicated COVID-19 cases had some diabetes related complications. However, to date, no study has been carried out to ascertain the extent of this due to the lack of accurate data.

> *“We (indicates health care professionals) need accurate data regarding COVID-19 tests and the infection rate to measure the severity to increase the service quality but hardly accuracy is being seen in existing data which if not considered would lead to upheaval” …(p29)*

##### Findings from Literature Search

Various newspapers reflected that Bangabondhu Sheikh Mujib Medical University (BSMMU) and the Bangladesh Diabetic Association (BDA) provided telemedicine that helped diabetes patients on varying levels of severity. Different pharmaceutical companies used social media platforms to provide diabetic care information and share their activities to support diabetic patients, with the government printing and disseminating posters regarding diabetes management. However, this campaign did not effectively reach the public as most patients were homebound, and did not see such posters hung in various hospital locations.

Information related to COVID-19 and test results were collected and distributed by the Institute of Epidemiology Disease Control and Research (IEDCR). However, they did not release data, specifically targeting diabetes or other co-morbidities associated with it.

#### Health Financing for Diabetes

##### Findings from Interviews and FGDs

Together with the frequent mention of need for sufficient budget allocation, the majority (89%) of the participants pointed to the underlying irregulation and corruption which is commonly defined as the “abuse of entrusted power for private gain” in heath sector [Table 6].

> *“Despite the need of more budget allocation aimed NCDs, corruption surged during COVID-19 which led to mismanagement and results a healthcare system more vulnerable” … (p32)*

Besides this, the failure of transportation systems with increased mandatory social distancing was mentioned by the respondents. Respondents also associated the increased prices of medicine, - Despite the country’s overall economic downturn brought on by the mandatory lockdown, - with it.

> *“Some 70% of the public’s total health costs have to be covered from pocket expenditures. Hence, without the government’s regulation of price gouging and inflation, costs increased, causing strain on the people” …(p35)*

Importantly, participants highlighted that the expense of insulin and other diabetic drugs increased well above the retail price at this time. Meanwhile, the innovative initiative of telemedicine has added to this financial strain, making it much more expensive than in-person visits along with reported difficulties in managing patients’ treatments (the patients could not properly explain the condition). Patients who belong to low-income group expressed their utter concern regarding livelihood than their health condition.

> “*When it has become really difficult for people like us to earn breads for survival in this COVID-19 situation, buying medicines for fancy disease like diabetes is beyond capacity” … (P40)*

##### Findings from Literature Search

While the government invested appreciable funds for the management of patients with COVID-19, most of it went towards the purchase of ventilators, incubator, masks, and personal protective equipment (PPE). The government also invested approximately USD 967 million for COVID-19 treatments and vaccines However, there was no specific allocation of funds specifically targeting diabetes management systems. The World Bank and ADB (Asian Development Bank) also provided an additional USD 145 million as COVID-19 emergency response finances, which again did not involve any separate funds for diabetes. Specifically, the Health Population Nutrition Sector Program (HPNSP) received no new public finances to help with the management of patients with diabetes in its annual budget review.

#### Health Resources for Diabetes

##### Findings from Interviews and FGDs

Most of the participants (service receivers and service providers) [Table 5; Table 6] expressed that an increasing fear of infection and governments’ mandates have made all these services shut down, creating an unimaginable crisis for the diabetes patients, especially in the first four months of the outbreak.

> *“Diabetes management requires many components and investigations, including blood glucose monitoring that were severely hindered during the imposed lockdown” …(p31)*

However, the study found that the co-payments associated with investigations made them accessible only for the patients who belong to the higher economic level. In contrast, patients in the lower socio-economic group were unable to go through such management as they relied on cheaper in-person systems.

In response to the question regarding the scope and necessity of research on COVID-19 and diabetes, patients mostly highlighted the deficiency in resources and shortage of accurate data.

> *“I together with some of my colleagues tried hard to carry out a study, investigating the relationship between diabetes and COVID-19, neither had we got financial support nor case related data, we delayed… (p24)*

##### Findings from Literature Search

The contribution of both the government and private pharmaceutical companies in resource mobilization for diabetes treatment research was disheartening. Meanwhile, physicians have suggested – in different health webinars that were documented by the newspapers - that by creating an online-based training platform for the frontline workers to enhance their skill and capacity, the diabetes related complications during COVID-19 can be handled.

#### Essential Medical Products and Technology for Diabetes

##### Findings from Interviews and FGDs

The physicians in focus group discussion felt that mobile phones and televisions can be used to spread information regarding diabetes management. However, due to the appreciable shortages of PPE, healthcare service workers hardly undertook echocardiograms, MRIs, ECGs and other essential tests during the COVID-19 pandemic as these involved close contact with patients.

> *“I believe, each one of us experienced a sense of dilemma for we too have feeling for family, at the meantime, it was hard to ignore the frightened eyes of our patients. The quality PPE seemed the only earthly rescuer then” … (p55)*

92% of participants responded not having the opportunity to buy the medications they are prescribed since the assumed price would be higher in their critical financial situation. Among the service receiver participants, 16% were not taking any antibiotics, insulin and other medications for diabetes control during lockdown.

##### Findings from Literature Search

Diabetes-related medical products became increasingly limited during lockdown. Endocrinologists could not be supplied quality PPE, and patients could not access various biological investigations needed to ensure a proper checkup to monitor possible complications. Bangladesh witnessed a 4% coronavirus mortality rate and 2.1% infection rate further constrained the treatment availability. An appreciable number of hand-sanitizer and hydroxychloroquine tablet were sold for the prevention of COVID-19 and Z-packs; however, treatments for DM and HTN remain undersold. However, poor slum and village people could not access appropriate medicines to manage their diabetes and associated complications due to the price. Cardiac diabetic complications can be properly investigated by echocardiograms, but Bangladesh has not developed a mobile testing system test for appropriate diagnosis. Hospitals lacked negative air pressure systems, so it was quite hard to prevent infection in the hospital premises.

##### Health Service Delivery for Diabetes

The severe impact of COVID-19 overburdened the the health system infrastructure in Bangladesh further impact on care delivery for patients with NCDs including diabetes. This included social-and-health service delivery

> *Mr. Rashid (pseudonym), a diabetic patient who lives in Dhaka, takes insulin two times a day to keep his glucose level in control and his daily routine involves morning and evening walk until lockdown has been imposed. He used to have a diet, including different types of fruits recommended by his physician. Mr. Rashid has no complain about life up to the period of lockdown when his neighborhood has suddenly detached from all basic services like other parts of the country*.
>
> *Mr. Rashid also confronted difficulties in finding his medication for diabetes in the local medical stores for most of those being imported. He further couldn’t continue his daily walk due to governments’ incumbent on people to stop the spread of COVID-19, and regular diet has been hampered for the disruption in food supply. Moreover, these triple triggers of service disruption created deep tension in the household of Mr. Rashid that he might be in danger of Hyperglycemia and Hypoglycemia*.

The mental pressure and physical turmoil of diabetic patients during lockdown in Bangladesh further exacerbated their care. Most of the participants (physicians) responded that those with diabetes had worse complications with COVID-19 infection, and maximum cases of COVID-19 related deaths had diabetes-related co-morbidities. This was also reported in the newspapers. Physicians reported that patients with COVID-19 and uncontrolled diabetes had a low recovery and higher death rate than those without this co-morbidity. Various diabetic association centers also had high physician infection rates, which in turn disturbed their services.

Participants shared some of their sufferings in the FDGs.

> *“We (participant and his wife) have no son and our only daughter is married; in this age (51-55) I can’t take the burdens of searching for medicines. She becomes extremely frightened and keeps asking me how we would cope with this kind of unprecedented situation” … (p44)*
>
> *“My parents diagnosed with COVID-19, and at the end my father (56-60) alone won the fight. Although my mother (46-50) was quite young, diabetes gave her a tough situation” … (p48)*

There were concerns with the knowledge gap in almost all of the participants of low-income and of low-educational status.

> *“I do not clearly understand what diabetes is or its management except that it is not curable. My rickshaw stand is near a hospital where various posters of health issues are hung on but due to being unable to read, do not understand much from them” … (p66)*

It is hard to measure the social and economic challenge caused by diabetes, which leads to both expensive maintenance and various life-threatening complications, most commonly stroke and neuropathy. However, governments’ public health service infrastructure was closed from March to July 2020. Diabetic association hospitals had many undiagnosed COVID-19 patients, and diabetic patients also did not share signs and symptoms with their physicians due to negative stigmas associated with COVID-19. Furthermore, Diabetes Association Hospitals’ ICU patients tested positive for COVID-19 were hospitalized, causing the spread of COVID-19 among health professionals that results a lockdown of ICU which in turn closed general services.

> *Ranu Begum (pseudonym), a working woman living in a well-sophisticated area of Dhaka, called her uncle, Mr. Zafor (pseudonym), and requested him to call her mother, Mrs. Roushon (pseudonym), to check on her as she was not picking up any calls from any of her children. She also informed about her mother being diabetes and other NCDs patient along with a high fever nowadays. Upon reaching Mr. Zafors’ call, Mrs. Roushon received and informed about her current situation. When Mr. Zafor asked her about not answering calls from her children, she mentioned a sorrow that has been very common during this pandemic hour*.

Social distancing and an underlying concept of fear regarding COVID-19 caused appreciable concerns with the mental health of people in such way that there were distances even between blood-relationship. The parents living with NCDs also under went the extra burden of stress and anxiety due to being ignored by their children.

> *Her sons live in the same building as her on different floors. Despite being informed that she has had a fever, they did not visit her even for once, rather insisted on taking over the phone, putting all the blame in the neck of the imposed lockdown. Hence, Mrs. Roushon became upset with her children’s response and stopped receiving their calls*.

The majority of the older adult participants with diabetes confronted both physical and psychological burden during the lockdown period, indicating stressed circumstance for patients with non-communicable diseases [Table 5]. Lockdown and social distancing on diabetes care had many harmful effects that may have been addressed by the implementation of robust diabetic patient guidelines.

## Discussion

The countrywide lockdown in response to COVID-19 appreciably impacted the supply chain for medicines, the closure of dispensaries, and a sudden increase in the price of insulin and other imported medicines making them unaffordable for patients with diabetes, particularly for those who live in low-economic setting. Alongside, the financial downturn of incomes with an increase in the number jobless-made people prioritize other areas instead of drug purchases.

Alongside, the treatment and management of chronic illnesses including diabetes has become even more complicated under government-mandated lockdown and quarantining. These measures including the closure of various services and clinics also affected access to treatment and drugs. There were concers about access and funding of health foods as well as space for exercise during lockdowns. with women often bearing the greater brunt of these limitations [96].

Bangladesh government’s expenditure in the health sector is the 3^rd^ biggest investment in the country which has a total of 62 district-level hospitals, 421 Upazilla health complexes, 1312 Union healthcare sub-centers, and Community Clinics. Additionally, three private sector hospitals and health-care centers are available for treating Diabetic patients. Moreover, there are dedicated diabetes-care units in government hospitals in Bangladesh. A recent study shows that, among the people living with diabetes, 9.8% are reported not taking any antidiabetic medication at all. Around 40.8% of the patients took insulin, 46.6% took metformin, 38.7% sulfonylurea, 38.7% any antihypertensive medication, and 14.2% took anti-lipids over the preceding three months [97]. Similarly, despite of the Health Population and Nutrition Sector Development Programme (HPNSDP) which has started transforming the health-care centers in favor of NCDs by extending the model to 70 Upazilas to provide diabetes and hypertension care, with an expectation to expand in 100 more Upazilas in the next 2 years. Alongside this, HPNSDP services were limited due to the country-wide lockdown, shortage of quality PPE, social stigma, and fear among the service providers. This is a concern going forward. Besides this, the lack of knowledge and evidence-based treatment guidelines for patients with COVID-19 during the pandemic has worsened the overall treatment continuity for diabetes care. Whilst the Ministry of Health and Family Welfare (MoHFW) in Bangladesh developed COVID-19 clinical management guidelines at the beginning of the pandemic, this did not include guidance regarding the care of patients with diabetes care; however, these were subsequently included in the later version of the guideline [98]. Alongside this, there was appreciable disruption in cardiac-care during pandemic. This though started to be addressed after several patients were refused such services, and this was reported in the media.

This limitation in service delivery during the pandemics resulted in a lack of satisfaction among people living with diabetes. The government’s recent initiative of providing training to 13,000 CHCPs about diabetes care can become a progressive step towards mitigating the gaps in the service and bring back satisfaction among service receivers. Additionally, the sustainable health care model for diabetes and general patients created by Diabetic Association of Bangladesh (BADAS) is playing an effective role in reducing the gaps brought about by the pandemic and ensuring quality service to the patients by treating more diabetic patients, with the expectation to increase the coverage to 75% patients by 2022 [99,100]. The Association currently manages over 100 institutions, ranging from small to large hospitals that provide primary, secondary, and tertiary care across all disciplines of diabetes. However, still the consequences of COVID-19 pandemic and it’s impact on the health system management is increasing the risks of diabetes in Bangladesh at a steady pace. Consequently, strengthening the healthcare system management is important to maximize the outcomes of existing diabetic care systems in Bangladesh.

The lack of sufficient PPE was a constant worry during the pandemic. A total of 8,042 healthcare professionals were affected by COVID-19 by October 01, 2020, and rising. Among them, 35% were doctors, 24% were nurses, and 41% were other healthcare staffs.

Human Resource Management is another essential area along with the infrastructures for the development of diabetes treatment. In Bangladesh, diabetes is widespread, but there is a gap in organized care for patients living with diabetes, and other chronic conditions. But often professional and civil society associations help fill this gap of caregiving, mostly taking no notice of policy level advocacy, and research. Thus, here arises another loophole that compliments the lack of patients’ knowledge, evidence-based policy change, and policy implementation aiming NCDs like diabetes. Whilst telemedicine services have been initiated and operated during the pandemic, there was dissatisfaction among both the service provider and receivers. This discontentment indicated to the necessity of management with all its concomitant area and capacity building training in health sector, with special focus on chronic diseases like diabetes along with COVID-19 diabetes management. In Bangladesh, there are diploma, post-graduation, and MD courses for diabetes. But in comparison, there are limited institutional opportunities for the root-level and technical workers to develop their skills. Expanding the scope of opportunities to the root-level can play a vital role in effective approach towards COVID-19 like possible pandemics in the future.

## Study Strengths

The strengths of the study include an in-depth look at factors affecting the health system response for the management of diabetes both from the patients and key healthcare professional’s viewpoint. The qualitative approaches grasp the economic contextualization of participants’ stance not as a matter of individual choice; but rather as the consequence of a politically-constructed financial direness; and the impact of economic barriers on the life of people living with diabetes and diabetes care that went beyond the issue of direct medical costs in a country like Bangladesh where the poverty and unemployment rates are high enough.

## Study Limitations

We are aware of a number of limitations with this study. Due to COVID-19, the study conducted the KIIs and FGDs through online platforms which limited our reach to those who had no internet access or supported devices. Besides, the unfamiliarity of online platforms including zoom and google meet made it difficult sometimes to communicate with the potential respondents who met study criteria. Additionally, the recruiting approach might have some sample bias when selecting respondents who live in remote or hard-to-reach areas. Despite these concerns we believe our findings are robust providing direction for the future.

## Priority for Future Research

There is an urgent need for research to determine the most effective control measures for diabetes, including the use of technology, in particular situations - such as the ICU, diabetic ketoacidosis, etc. - during such pandemic hours. Besides, future study would be best served by examining the need for diabetes health support at this stage of the pandemic and by using similar or identical questions throughout all iterations to learn more about how the health system will function, what information patients may be seeking, how patients find or provide support, and whether patients feel empowered to seek additional help from a healthcare professional when necessary.

## Implication for Policy and Clinical Practice

The study should help the relevant authority to better bring their expertise together with that of the patients to craft more personalized and effective diabetes management strategies. The study found lack of expertise among people living with diabetes regarding accessing information and resource, hence providing high-quality structured training in local language to support the skills of diabetes self-management is strongly suggested. The majority of persons with diabetes receive care from primary health care services, hence it is crucial to strengthen the structural capabilities of primary diabetes care practices. Besides, the results suggest that Bangladesh needs to significantly enhance diabetes detection and treatment, particularly among underprivileged groups. And, these can be addressed by reorganizing the healthcare system in accordance with disease burden; the government should prioritize NCDs, notably the prevention and control of diabetes; initiating advocacy activities and implementing universal health and health financing system to ensure access and affordable care for all.

## Conclusion

The healthcare system of Bangladesh clearly lacks attention to the growing burden of chronic non-communicable diseases. Besides, the low SES barely gives opportunity to the people living with diabetes to know the reasons behind their health complexities due to non-compliance and non-adherences. Further strengthening of the health system response is ineluctable to enable for effective confrontation of challenges posed by NCDs. Besides, patient education should be given extra emphasize, so as giving the modern world. Both the privileged or unprivileged population should be brought under the arena of compliance through patient education, at least by health. Telemedicine service should be strengthened to ensure easy access to the services for diabetic patients. Moreover, appropriate measures should be taken to make the necessary medication available for the people living with diabetes along with popularizing the testing tools and technology among the mass people. Finally, necessary measures should be taken to motivate the people to follow a healthy lifestyle and raise awareness about the potential risk factors of Diabetes Mellitus. Taking these measures can gradually bring some change in the healthcare system of diabetes treatment in Bangladesh.

## Data Availability

All data produced in the present study are available upon reasonable request to the authors

## Ethical Approval

The study protocol was approved by the authorities of Institutional Reviewed Board (IRB) of Eminence Associates for Social Development containing number (Emi/IRB/Oct/2010011001). The study carried out all the methods in accordance with institutional, relevant, national or international guidelines and regulations.

## Consent to Publication

‘NOT APPLICABLE’

## Availability of Data and Materials

The data that support the findings of this study are available from Eminence Associates for Social Development but restrictions are applied to the availability of these data, which were used under license (Emi/IRB/Oct/2010011001) for the current study, and so are not publicly available. Data are however available from the authors upon reasonable request and with permission of Eminence Associates for Social Development.

## Authors’ Contributions

The correspondent author SHT and his team members TI, KFS, SA, DF, UFS vi, SMSI and ZS and AH have equally participated (Data analysis, Report writing, Literature review) in this study.

## Acknowledgements

The authors are grateful to the anonymous referees of the Journal for their beneficial recommendations for developing the excellence of the paper. The standard disclaimers apply.

